# Cross-sectional study to assess the efficacy of SARS-CoV-2 vaccination in patients with monoclonal gammopathies

**DOI:** 10.1101/2022.01.19.22269531

**Authors:** Eugenia Abella, Macedonia Trigueros, Edwards Pradenas, Francisco Muñoz-Lopez, Francesc Garcia-Pallarols, Randa Ben Azaiz Ben Lahsen, Benjamin Trinité, Victor Urrea, Silvia Marfil, Carla Rovirosa, Teresa Puig, Eulàlia Grau, Anna Chamorro, Ruth Toledo, Marta Font, Dolors Palacín, Francesc Lopez-Segui, Jorge Carrillo, Nuria Prat, Lourdes Mateu, Bonaventura Clotet, Julià Blanco, Marta Massanella, on behalf of the VAC-COV-GM-HMAR, KING cohort extension and CoronAVI@S studies

## Abstract

SARS-CoV-2 vaccination is the most effective strategy to protect patients with haematologic malignancies against severe COVID-19, but primary vaccine responses are less effective in this population. Here, we characterized the humoral responses following 3 months after mRNA-based vaccines in patients at different stages of the same plasma cell diseases, including monoclonal gammopathy of undetermined significance (MGUS), smoldering multiple myeloma (SMM) and multiple myeloma on first line therapy (MM), compared to a healthy control population matched by sex and age. We observed that plasmas from uninfected MM patients after 3 months post-vaccine have lower SARS-CoV-2 specific IgG and IgA antibodies and decreased neutralization capacity compared with MGUS and SMM individuals, and a group of healthy controls. Importantly, we detected significantly higher plasma neutralization capacity in MM individuals who recovered from COVID-19 compared to their uninfected counterparts, highlighting that hybrid immunity elicit stronger immune responses even in this immunocompromised population. In contrast to MM group, no differences in the vaccine-induced humoral response were observed between uninfected MGUS, SMM and healthy individuals. In conclusion, a booster vaccine dose is recommended in uninfected MM patients to develop an adequate and effective humoral response to SARS-CoV-2 vaccine.

## Introduction

Cancer patients present substantial immune impairment induced by the tumour itself or by their treatment, and therefore they are at increased risk for infections and infection-related mortality. In fact, SARS-CoV-2 infection in non-vaccinated cancer patients with immunosuppression has been associated with significantly higher morbidity and mortality rates [1], especially those patients with haematological neoplasms [2]. Patients with plasma cell dyscrasias, such as multiple myeloma, are associated with immunosuppression and at increased risk of infections due to various circumstances: active disease status, decrease in non-clonal immunoglobulins (immunoparesis) and impairment of cellular immunity, but also can be affected by comorbidities and older age [3]. In addition, new approaches with proteasome inhibitors, immunomodulators, monoclonal antibodies and CAR T cells may also exacerbate this immune dysfunction [4], putting these individuals at higher risk of any infection. Indeed, at least 80% of multiple myeloma patients required hospital admission after SARS-CoV-2 infection [5], and more than 30% died because of COVID-19 [4]. SARS-CoV-2 vaccination is the most effective strategy to protect these vulnerable population against severe COVID-19, however previous studies showed reduced humoral responses against several vaccines (i.e., pneumococci, staphylococcal alpha toxin, tetanus, among others) [6].

Given their greater susceptibility to severe COVID-19 and lower vaccine-induced immune responses, patients with monoclonal gammopathies are a high-priority group for vaccination to mitigate COVID-19 related morbidity and mortality [7] and further characterization of their immunity generated are required. Current guidelines recommend SARS-CoV-2 vaccination of all patients with monoclonal gammopathy of undetermined significance (MGUS), smoldering multiple myeloma (SMM) and multiple myeloma on therapy (MM) [8]. It is important to determine the protection achieved with the standard 2 doses vaccination to adapt the vaccination calendar to their immune needs and prioritize a third dose administration, if necessary. Published data about SARS-CoV-2 vaccination efficacy refers mainly to patients with MM and, to a lesser extent, with SMM [9–12]. Data in patients with MGUS are scarce and requires further investigation.

The aim of this study was to evaluate the COVID-19 vaccine humoral responses differ in patients with monoclonal gammopathies, including MGUS, SMM and MM, compared to a healthy control population, to adjust the vaccination calendar to the subtype of disease.

## Material and methods

### Study population

A cross-sectional study (VAC-COV-GM-HMAR) was conducted in patients with monoclonal gammopathies at Hospital del Mar (Barcelona, Spain) to assess the efficacy of vaccination against SARS-CoV-2, after having received the complete vaccination schedule dose according to BNT162b2 (Pfizer-BioNTech) and mRNA-1273 COVID-19 (Moderna) schedules. Fifty-nine patients suffering monoclonal gammopathies consecutively visited during 2 months at the outpatient haematological consult were eligible to entry into the study. Treated patients received only one line of therapy before vaccination. Blood samples were obtained a median of 3.9 IQR [2.2-5] months after complete vaccination schedule.

Results were compared to an uninfected control group, which included 36 individuals without haematological malignancies, belonging to the *King cohort extension (N=22)* and CoronAVI@S (N=14) studies. Post-vaccine samples were selected among individuals vaccinated also with BNT16b2 and mRNA-1273 COVID-19.

The VAC-COV-GM-HMAR, *King cohort extension* and CoronAVI@S studies were approved by the Ethics Committee Boards from the Hospital del Mar (HMAR), the Hospital Universitari Germans Trias i Pujol (HUGTIP) and the Institut Universitari d’Investigació en Atenció Primària (IDIAP) respectively (HMAR/2021/9913/I, HUGTiP/PI-20-217 and IDIAP/20-116P) and were conducted in accordance with the Declaration of Helsinki. All patients provided written informed consent before starting the study.

### Determination of anti-SARS-CoV-2 antibodies

The presence of anti-SARS-CoV-2 antibodies against Spike S2 Subunit+ Spike protein receptor binding domain (S2+RBD) or Nucleocapside protein (NP) in plasma samples was evaluated using an in-house developed sandwich-ELISA, as previously described [13]. The specific signal for each antigen was calculated after subtracting the background signal obtained for each sample in antigen-free wells. Values are plotted into the standard curve. Standard curve was calculated by plotting and fitting the log of standard dilution (in arbitrary units) vs. response to a 4-parameter equation in Prism 8.4.3 (GraphPad Software). See supplemental material.

### Pseudovirus neutralization assay

Neutralization assay was performed using SARS-CoV-2.SctΔ19 WH1 and B.1.617.2/Delta pseudovirus as previously described [14,15]. The lower limit of detection was 60 and the upper limit was 14,580 (reciprocal dilution). The ID50 (the reciprocal dilution inhibiting 50% of the infection) was calculated by plotting and fitting the log of plasma dilution vs. response to a 4-parameter equation in Prism 8.4.3 (GraphPad Software). See supplemental material.

### Statistical analysis

Quantitative variables were compared using the Mann–Whitney test and proportions using the chi-squared test for comparison between groups, and nonparametric Kruskal–Wallis test for comparison between groups. The impact of each variable to the levels of specific SARS-CoV-2 IgG or IgA antibodies and neutralization capacity was assessed via linear regression models including all patients with monoclonal gammopathies and the control groups, using the latter as reference. A multivariate model was constructed based on significant variables on the univariate analysis with an inclusion criterion of *p* < 0.1. Statistical analyses were performed with Prism 9.1.2 (GraphPad Software) and R (4.1.2). Statistical significance was determined when *p* ≤ 0.05.

## Results

### Characteristics of the individuals with monoclonal gammopathies

We recruited 59 patients with monoclonal gammopathies, who were sub-grouped according to their diagnosis (MGUS, SMM and MM). We took advantage of the national vaccination plan in cancer patients to evaluate the immune responses generated by the BNT16b2 or mRNA-1273 COVID-19. Six patients were excluded because they had received AstraZeneca or Janssen vaccination (Supplementary Figure 1). All samples were collected after a median of 3.9 IQR [2.2-5] months from complete schedule of vaccine administration (two doses). Additional characteristics of patients and their gammopathies are presented in Table 1.

**Table 1.** Patient and disease characteristics.

|  | MGUS |  | *P-value*<br>*between*<br>*infected and*<br>*uninfected<sup>†</sup>* | SMM | MM |  | *P-value*<br>*between*<br>*infected*<br>*and*<br>*uninfected<sup>†</sup>* |
| --- | --- | --- | --- | --- | --- | --- | --- |
|  | Uninfected | Infected |  | Uninfected | Uninfected | Infected |  |
|  | N=15 | N=2 |  | N=22 | N=10 | N=4 |  |
| Female, n (%) | 9 (60) | 2 (100) | 0.51 | 15 (68) | 5 (50) | 4 (100) | 0.22 |
| Age, years, median [IQR] | 67 [51-83] | 78 [77-78] | 0.51 | 73 [63-80] | 74 [68-79] | 53 [45-83] | 0.23 |
| Gammopathy subtype, n (%) |  |  |  |  |  |  |  |
| IgG (Kappa or Lambda) | 8 (53) | 1 (50) |  | 14 (64) | 6 (60) | 3 (75) |  |
| IgA (Kappa or Lambda) | 3 (20) | 1 (50) |  | 5 (23) | 2 (20) | 0 (0) |  |
| IgM (Kappa or Lambda) | 4 (27) | 0 (0) |  | 2 (9) | 0 (0) | 0 (0) |  |
| Bence Jones Kappa | 0 (0) | 0 (0) |  | 1 (5) | 1 (10) | 1 (25) |  |
| Bence Jones Lambda | 0 (0) | 0 (0) |  | 0 (0) | 1 (10) | 0 (0) |  |
| Immunoparesis, n (%) | 6 (35) | 1 (50) | 1.0 | 17 (77) | 9 (90) | 3 (75) | 0.10 |
| Total Lymphocytes (x10<sup>3</sup>),<br>cells/μl, median [IQR] | 2.5 [2.0-3.0] | 1.8 [1.3-2.4] | 0.37 | 1.8 [1.6-2.4] | 1.5 [0.8-2.3] | 1.8 [1.2-2.3] | 0.84 |
| Lymphopenia (<10<sup>3</sup> cells/μl), n (%) | 0 (0) | 0 (0) | 1 | 1 (5) | 3 (30) | 0 (0) | 0.5 |
| Treatment |  |  |  |  |  |  |  |
| No Treatment, n (%) | 17 (100) | 2 (100) |  | 22 (100) | 0 (0) | 0 (0) |  |
| VRD | 0 (0) | 0 (0) |  | 0 (0) | 2 (20) | 3 (75) |  |
| VRD+autoTPH | 0 (0) | 0 (0) |  | 0 (0) | 1 (10) | 0 (0) |  |
| Daratumumab in combination | 0 (0) | 0 (0) |  | 0 (0) | 4 (40) | 0 (0) |  |
| KRD | 0 (0) | 0 (0) |  | 0 (0) | 1 (10) | 0 (0) |  |
| KRD+auto-HSCT | 0 (0) | 0 (0) |  | 0 (0) | 1 (10) | 0 (0) |  |
| VTD+auto-HSCT | 0 (0) | 0 (0) |  | 0 (0) | 1 (10) | 0 (0) |  |
| Prednisone |  |  |  |  | 0 (0) | 1 (25) |  |
| Detectable anti-NP SARS-CoV-2 IgG, n (%) | 0 (0) | 2 (100) |  | 0 (0) | 0 (0) | 4 (100) |  |
| Vaccine, n (%) |  |  |  |  |  |  | 0.58 |
| Pfizer | 13 (87) | 2 (100) | 1.0 | 9 (41) | 3 (30) | 2 (50) |  |
| Moderna | 2 (13) | 0 (0) |  | 13 (59) | 7 (70) | 2 (50) |  |
| Time from complete vaccine dose to extraction, Days.Median [IQR] | 97 [42-144] | 65 [64-65] | 0.36 | 123 [67-158] | 80 [62-153] | 111 [77-177] | 0.81 |
| Range | 23-166 | 64-65 |  | 28 -167 | 33-156 | 77-188 |  |
| Comorbidities, n (%) |  |  |  |  |  |  |  |
| Hypertension | 6 (40) | 2 (100) |  | 8 (36) | 7 (70) | 2 (50) |  |
| Autoimmune disease | 0 (0) | 0 (0) |  | 0 (0) | 0 (0) | 0 (0) |  |
| Allergies | 1 (7) | 0 (0) |  | 4 (18) | 2 (14) | 0 (0) |  |
| Diabetes | 2 (13) | 0 (0) |  | 4 (18) | 2 (14) | 0 (0) |  |

<sup>†</sup>P-values obtained from Fisher exact test for categorical variables and Mann Withney for continuous variables.
HSCT: hematopoietic stem-cell transplantation; VRD: bortezomib, lenalidomide, dexamethasone; KRD: carfilzomib, lenalidomide, dexamethasone; VTD: bortezomib, thalidomide, dexamethasone

### Impact of hybrid immunity in immune responses to vaccine in patients with monoclonal gammopathies

Despite none of the participants included in this study had a positive SARS-CoV-2 PCR, we evaluated the presence of anti-NP SARS-CoV-2 specific IgG antibodies in all plasma samples to verify previous SARS-CoV-2 infection. Unexpectedly, we identified 2/17 (12%) and 4/14 (29%) patients from MGUS and MM groups, respectively, were previously infected by SARS-CoV-2. Since the combination of natural infection and vaccine-generated immunity elicit higher specific immune responses, uninfected and previously infected subjects suffering from monoclonal gammopathies were analysed separately. Despite the limited number of infected patients in our cohort, we found a statistical increase of circulating specific SARS-CoV-2 IgG and IgA antibodies against S2+RBD in previously infected MM patients compared to the uninfected counterparts (*p*=0.003 and *p*=0.04, respectively, Figure 1A and B). In contrast, a tendency of a reduction of specific SARS-CoV-2 IgG antibodies was found in infected MGUS patients compared to the uninfected group (*p*=0.09), while no differences were observed in the levels of specific IgA. Low levels of specific IgM were detected in all groups (Figure 1C). Finally, plasma from infected MM patients showed statistically increased levels of neutralization compared to their uninfected counterparts (*p*=0.01, Figure 1D), while no differences were observed between MGUS infected and uninfected groups.

**Figure 1.**
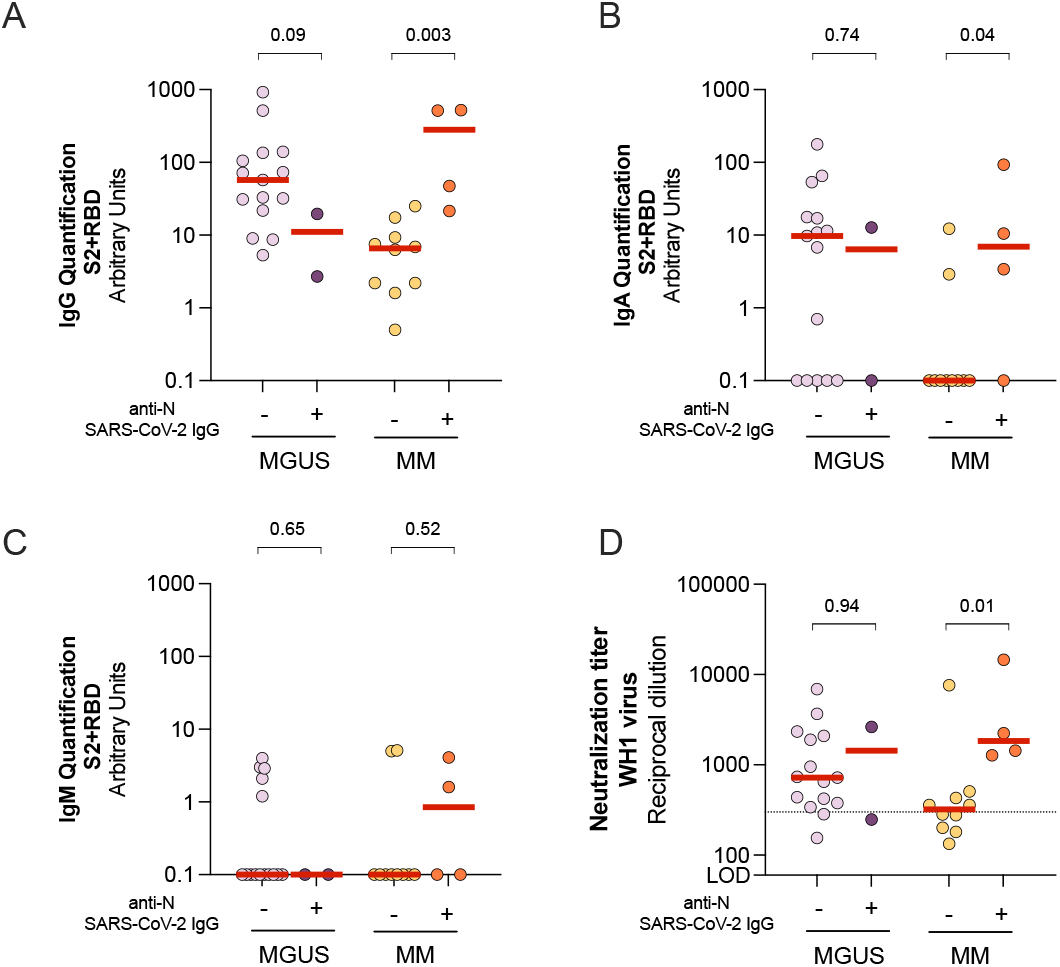
Comparison of humoral between uninfected and infected individuals suffering from monoclonal gammopathies. Levels of specific SARS-CoV-2 immunoglobulins IgG (Panel A), IgA (Panel B) and IgM (Panel C) against S2+RBD proteins quantified in plasma from uninfected and infected patients by ELISA. <u>Panel D:</u> Neutralizing activity against WH1 virus after three months of vaccine administration in infected and uninfected patients. Dotted line indicates the clinically relevant neutralization titer. In all panels, median values are indicated and p-values were obtained from Mann–Whitney test for comparison between groups.

### Humoral responses after 3 months from vaccination of uninfected individuals

Due to these differences in immune response to vaccine, we excluded COVID-19 recovered patients from our analysis to determine specifically the vaccine-generated immunity in patients with monoclonal gammopathies. An uninfected control group (CG) was included in our analysis as a reference group (Table 2) matched by age, sex and time after complete vaccination schedule.

**Table 2.** Characteristics from Uninfected patients and CG

|  | **MGUS** | **SMM** | **MM** | *P-value*<br>*between*<br>*monoclonal*<br>*gammopathies<sup>1</sup>* | **CG** | *P-value*<br>*between*<br>*all groups* |
| --- | --- | --- | --- | --- | --- | --- |
|  | **N=15** | **N=22** | **N=10** |  | **N=36** |  |
| Female, n (%) | 9 (60) | 15 (68) | 5 (50) | 0.61 | 21 (58) | 0.78 |
| Age, years, median [IQR] | 67 [51-83] | 73 [63-80] | 74 [68-79] | 0.56 | 62 [48-75] | 0.61 |
| Gammopathy subtype, n (%) |  |  |  |  |  |  |
| IgG (Kappa or Lambda) | 8 (53) | 14 (64) | 6 (60) |  | NA |  |
| IgA (Kappa or Lambda) | 3 (20) | 5 (23) | 2 (20) |  | NA |  |
| IgM (Kappa or Lambda) | 4 (27) | 2 (9) | 0 (0) |  | NA |  |
| Bence Jones Kappa | 0 (0) | 1 (5) | 1 (10) |  | NA |  |
| Bence Jones Lambda | 0 (0) | 0 (0) | 1 (10) |  | NA |  |
| Immunoparesis, n (%) | 6 (35) | 17 (77) | 9 (90) | **0.02** | NA |  |
| Total Lymphocytes ($\times 10^6$),<br>cells/μl, median [IQR] | 2.5 [2.0-3.0] | 1.8 [1.6-2.4] | 1.5 [0.8-2.3] | **0.006** | NA | |
| Lymphopenia ($<10^3$ cells/μl), n (%) | 0 (0) | 1 (5) | 3 (30) | **0.03** | NA | |
| Treatment |  |  |  |  |  |  |
| No Treatment, n (%) | 17 (100) | 22 (100) | 0 (0) |  | NA |  |
| VRD | 0 (0) | 0 (0) | 2 (20) |  |  |  |
| VRD+auto-HSCT | 0 (0) | 0 (0) | 1 (10) |  |  |  |
| Daratumumab in combination | 0 (0) | 0 (0) | 4 (40) |  |  |  |
| KRD | 0 (0) | 0 (0) | 1 (10) |  |  |  |
| KRD+auto-HSCT | 0 (0) | 0 (0) | 1 (10) |  |  |  |
| VTD+auto HSCT | 0 (0) | 0 (0) | 1 (10) |  |  |  |
| Vaccine, n (%) |  |  |  | **0.006** |  | **0.0004** |
| Pfizer | 13 (87) | 9 (41) | 3 (30) |  | 29 (81) |  |
| Moderna | 2 (13) | 13 (59) | 7 (70) |  | 7 (32) |  |
| Time from complete vaccine dose to extraction, Days |  |  |  |  |  |  |
| Median [IQR] | 97 [42-144] | 123 [67-158] | 80 [62-153] | 0.59 | 101 [90-111] | 0.44 |
| Range | 23-166 | 28 -167 | 33-156 |  | 33-184 |  |
| Comorbidities, n (%) |  |  |  |  |  |  |
| Hypertension | 6 (40) | 8 (36) | 7 (70) |  | 10 (28) | 0.11 |
| Autoimmune disease | 0 (0) | 0 (0) | 0 (0) |  | 0 (0) | 1.0 |
| Allergies | 1 (7) | 4 (18) | 2 (14) |  | 4 (11) | 1.0 |
| Diabetes | 2 (13) | 4 (18) | 2 (14) |  | 7 (19) | 1.0 |

<sup>1</sup>P-values obtained from Fisher exact test for categorical variables and Kruskal–Wallis for continuous variables
HSCT: hematopoietic stem-cell transplantation; VRD: bortezomib, lenalidomide, dexamethasone; KRD: carfilzomib, lenalidomide, dexamethasone; VTD: bortezomib, thalidomide, dexamethasone

All individuals seroconverted after COVID-19 vaccination, however there were statistically significant differences among groups in the levels of specific SARS-CoV-2 IgG antibodies (Kruskal-Wallis *p*=0.002, Figure 2A). Patients with MM showed significantly lower levels of SARS-CoV-2 specific IgG antibodies compared to all groups (p<0.006 in all cases, Figure 2A), while no differences were observed between MGUS and SMM with control group. Lower levels of specific SARS-CoV-2 IgA antibodies were detected and 33% (5/15), 23% (5/22) and 80% (8/10) of MGUS, SMM and MM did not develop any specific IgA antibodies, respectively (Fisher exact test *p*=0.008, Figure 2B). Again, MM showed significantly lower levels of SARS-CoV-2 specific IgA antibodies compared to CG and MGUS (*p*=0.002 and *p*=0.04, respectively) and a tendency compared to SMM (*p*=0.08). SARS-CoV-2 specific IgM antibodies were almost undetectable in all samples, including healthy CG (Figure 2C). Similarly to the serology results, patients with MM showed a tendency for lower neutralization capacity compared to CG (*p*=0.08, 321 [196-449] and 770 [422-1554] neutralization titer, respectively), while no differences in neutralization levels between MGUS or SMM with CG were observed (Figure 2D). Considering the cut-off of 300 neutralization titer that could provide a relevant level of protection, we found that 5/10 (50%) of MM patients did not reach this neutralization threshold in comparison of the 5/36 (14%) of the CG individuals (Fisher exact test, *p*=0.03). In contrast, patients suffering from MGUS or SMM showed comparable percentages of patients below the relevant neutralization activity compared to the CG group (2/15 [13%] and 3/22 [14%], respectively). We then estimated the proportion of effective neutralizing antibodies among the total SARS-CoV-2 specific IgG, calculated as the ratio of plasma neutralization titer to total SARS-CoV-2 IgG antibodies. We observed an increase of this ratio in patients with MM compared to other groups (Figure 2E, *p*=0.02), suggesting that despite the low levels of specific SARS-CoV-2 antibodies in this population, these antibodies showed to be functional.

**Figure 2:**
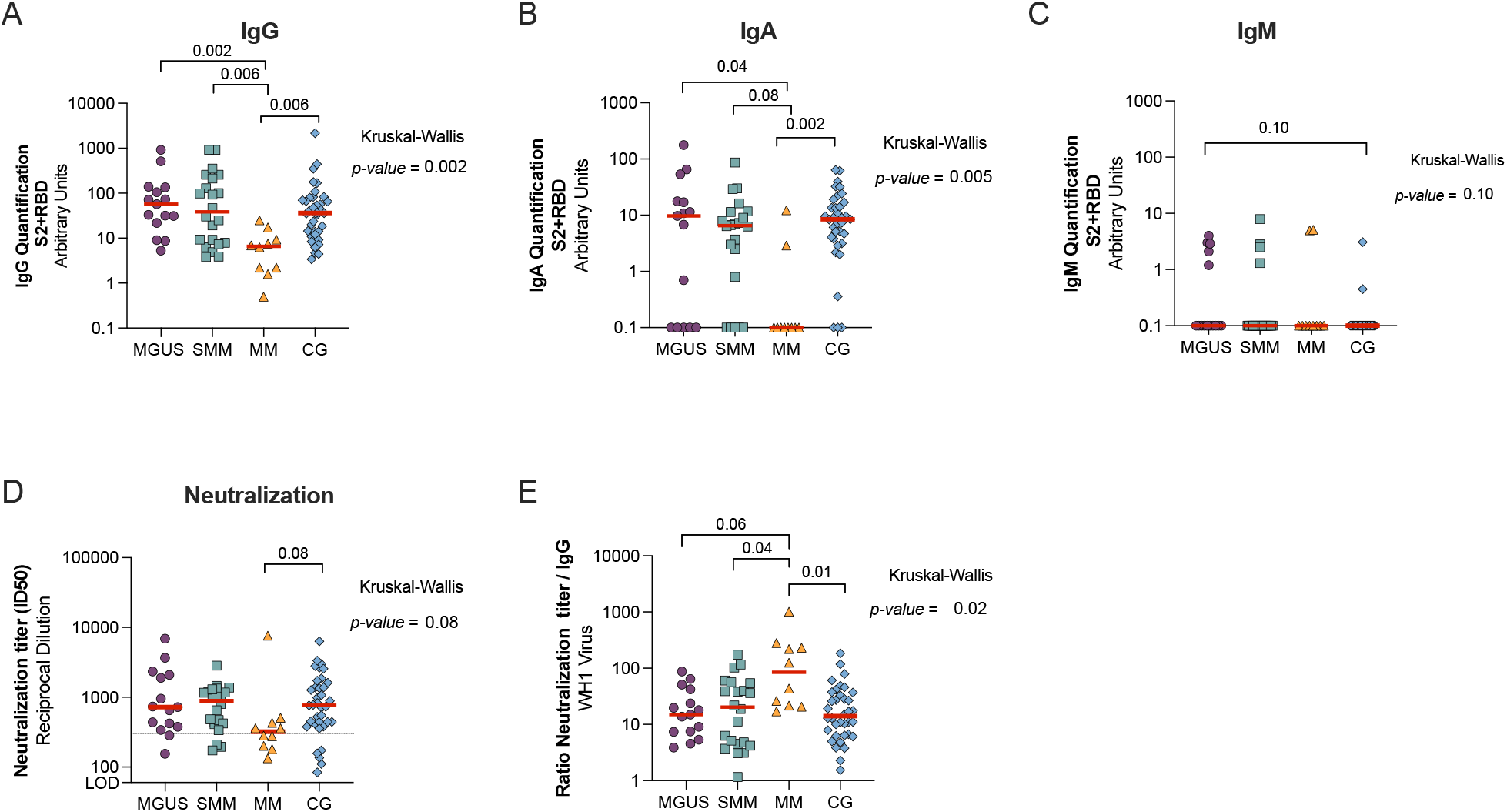
Comparison of humoral response after 3 months from mRNA vaccination in uninfected patients with monoclonal gammopathies compared to a control group (CG). Levels of specific SARS-CoV-2 immunoglobulins IgG (Panel A), IgA (Panel B) and IgM (Panel C) against S2+RBD proteins quantified and neutralizing activity against WH1 (Panel D) from MGUS, SMM and MM patients and a control group (CG) after three months of vaccine administration. Dotted line indicates the clinically relevant neutralization titer (panel D). Panel E: Ratio of plasma neutralization titer per total SARS-CoV-2 IgG antibodies. Median values are indicated; P-values were obtained from Kruskal–Wallis test for comparison between groups.

Since specific treatments received by MM group could cause immune dysfunction impairing immune response to COVID-19 vaccines, we first compared the levels of specific SARS-CoV-2 IgG antibodies and neutralization capacity in MM patients receiving or not Daratumumab (anti-CD38 therapy). No differences were found between groups (Supplementary Figure 2A and B). Similar results were found when we compared MM individuals who underwent autologous hematopoietic cell transplantation (auto-HSCT) *vs*. no auto-HSCT (Supplementary Figure 2 C and D). Despite the limited number of patients, these results suggest that these treatments might not the main cause of the immune dysfunction observed in these individuals.

### Predictive factors for specific SARS-CoV-2 humoral responses

There was a positive correlation between SARS-CoV-2 specific IgG and IgA antibody response and the neutralization titer (r=0.60 *p*<0.0001, and r=0.54 *p*<0.0001, respectively, Figure 3 A, B and C). On the other hand, we found a negative correlation between the neutralization capacity and the days post-complete vaccine scheduled (r=–0.48, *p*<0.0001) and age (r=–0.27, *p*<0.0001). In addition, age was also negatively correlated to the levels of circulating SARS-CoV-2 specific IgG and IgA antibodies (r=-0.37, p<0.0001 and r=-0.34, *p*=0.002, respectively). Due to the potential impact of different parameters in the humoral response described in the literature and found in our cohort, we performed linear regression models for the levels of specific SARS-CoV-2 IgG and IgA antibodies and neutralization capacity for the variables: group (MGUS, SMM, MM and CG [reference]), age, sex, immunoparesis (yes/no), vaccine (Pfizer/Moderna), days post complete vaccine schedule and sub-type of gammopathy (IgG, IgA, IgM, light chain). In the univariate analysis, we found that MM group, age and immunoparesis were significantly associated with lower levels of circulating SARS-CoV-2 specific IgG antibodies (Table 3A). In the multivariate analysis, only MM group and age were negative predictors of lower levels of SARS-CoV-2 IgG antibodies. Similarly, MM group, age, IgG gammopathy and immunoparesis were significant predictive factors for lower levels of circulating SARS-CoV-2 specific IgA antibodies in the univariate analysis, and only age remained significant in the multivariate analysis. Finally, MM group, age, immunoparesis and days post complete schedule were significant predictive factors for lower neutralization capacity whereas Moderna vaccine had a tendency to higher neutralization levels (Table 3C). The multivariate analysis revealed that all these parameters remained significant predictors, except for immunoparesis.

**Figure 3:**
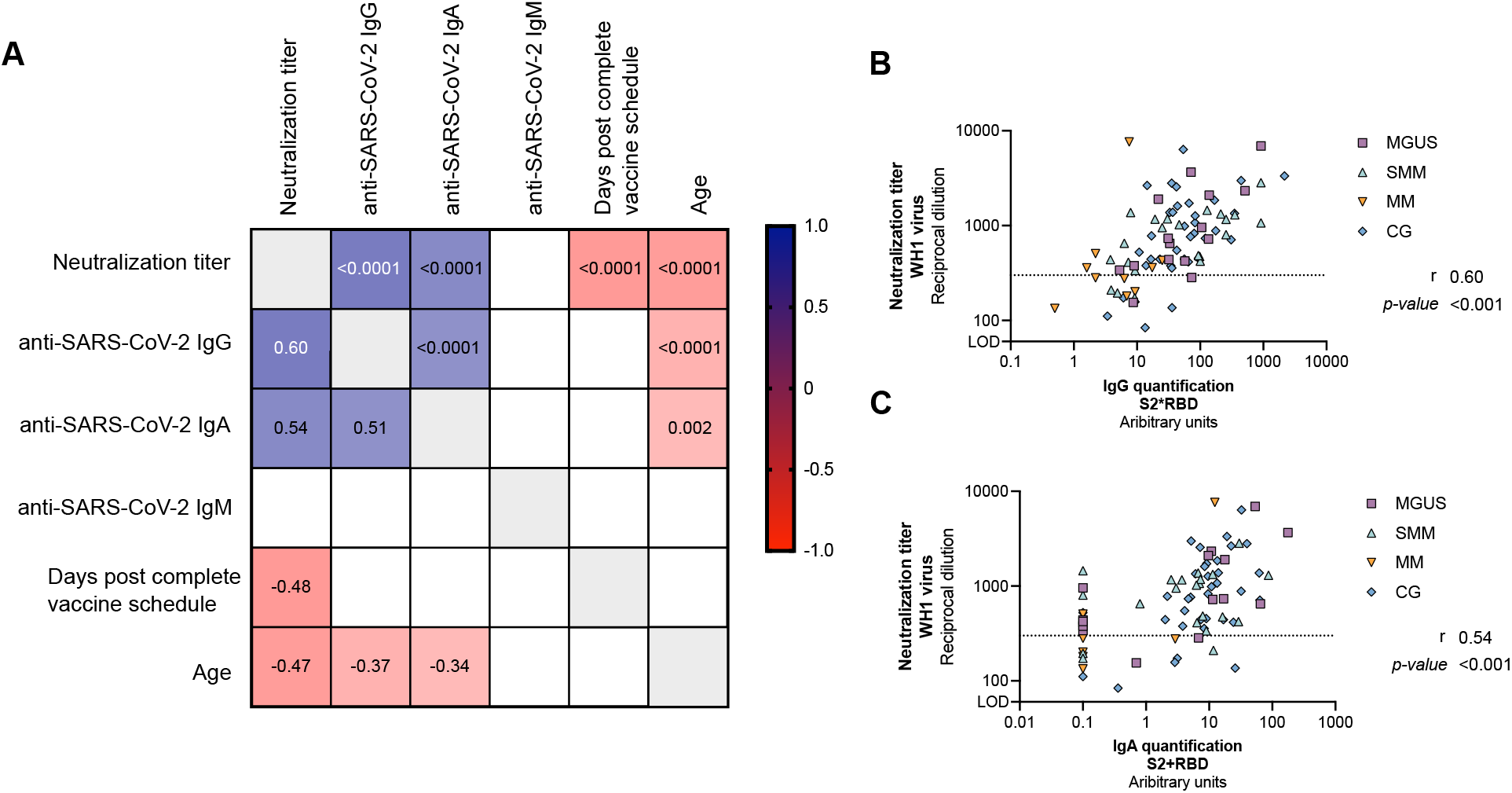
Correlations between variables. Panel A: Correlation matrix of relevant continuous variables including MGUS, SMM, MM and CG participants. Spearman coefficients are indicated in the lower part of the panel, while p-values in the upper part. Only significant correlations are plotted (p<0.05). Positive correlations are shown in blue, while negative in red. Detail of the correlation between the levels of SARS-CoV-2 specific IgG (Panel B) and IgA (Panel C) antibodies and neutralization capacity. Each symbol represents a participant, and are color-coding according to their disease group (MGUS: purple square, SMM: upper triangle turquoise, MM: lower triangle orange and CG: blue diamond). Correlation coefficient and p-values were obtained from Spearman correlation.

**Table 3:** Univariate and Multivariate analysis

**A. Levels of SARS-CoV-2 specific IgG**
|  | Univariate |  |  | Multivariate |  |  |
| --- | --- | --- | --- | --- | --- | --- |
|  | Estimated | St error | P-value | Estimated | St error | P-value |
| Group [Reference CG] |  |  |  |  |  |  |
| MGUS | 0.041 | 0.193 | 0.832 | 0.152 | 0.197 | 0.444 |
| SMM | -0.056 | 0.170 | 0.744 | 0.219 | 0.230 | 0.343 |
| MM | -1.016 | 0.225 | **<0.0001** | -0.686 | 0.287 | **0.019** |
| Sex (Woman) | -0.119 | 0.156 | 0.446 |  |  |  |
| Age | -0.016 | 0.005 | **0.002** | -0.013 | 0.005 | **0.007** |
| Gammopathy subtype |  |  |  |  |  |  |
| IgG (Kappa or Lambda) | -0.337 | 0.176 | 0.059 |  |  |  |
| IgA (Kappa or Lambda) | 0.129 | 0.250 | 0.607 |  |  |  |
| IgM (Kappa or Lambda) | -0.281 | 0.308 | 0.364 |  |  |  |
| Bence Jones Kappa | -0.242 | 0.508 | 0.635 |  |  |  |
| Bence Jones Lambda | -0.446 | 0.708 | 0.530 |  |  |  |
| Immunoparesis | -0.438 | 0.153 | **0.005** | -0.245 | 0.213 | 0.253 |
| Days post-complete schedule | -0.003 | 0.002 | 0.117 |  |  |  |
| Vaccine kind (Moderna) | 0.157 | 0.161 | 0.335 |  |  |  |

B. Levels of SARS-CoV-2 specific IgA
|  | Univariate |  |  | Multivariate |  |  |
| --- | --- | --- | --- | --- | --- | --- |
|  | Estimated | St error | P-value | Estimated | St error | P-value |
| Group [Reference CG] |  |  |  |  |  |  |
| MGUS | -0.089 | 0.176 | *0.617* | 0.038 | 0.181 | 0.834 |
| SMM | -0.183 | 0.155 | *0.242* | 0.125 | 0.211 | 0.557 |
| MM | -0.735 | 0.205 | **<0.001** | -0.370 | 0.263 | *0.164* |
| Sex (Woman) | -0.080 | 0.135 | *0.555* |  |  |  |
| Age | -0.012 | 0.004 | **0.004** | -0.009 | 0.004 | **0.030** |
| Gammopathy subtype |  |  |  |  |  |  |
| IgG (Kappa or Lambda) | -0.316 | 0.151 | **0.040** |  |  |  |
| IgA (Kappa or Lambda) | -0.087 | 0.214 | *0.686* |  |  |  |
| IgM (Kappa or Lambda) | -0.374 | 0.264 | *0.161* |  |  |  |
| Bence Jones Kappa | 0.079 | 0.436 | *0.856* |  |  |  |
| Bence Jones Lambda | -0.890 | 0.608 | *0.147* |  |  |  |
| Immunoparesis (yes) | -0.436 | 0.130 | **0.001** | -0.317 | 0.195 | *0.109* |
| Days post-complete schedule | -0.002 | 0.002 | *0.280* |  |  |  |
| Vaccine kind (Moderna) | 0.202 | 0.139 | *0.151* |  |  |  |

C. Neutralization Capacity
|  | Estimated | Univariate<br>St error | P-value | Estimated | Multivariate<br>St error | P-value |
| --- | --- | --- | --- | --- | --- | --- |
| Group [Reference CG] |  |  |  |  |  |  |
| MGUS | 0.010 | 0.129 | *0.941* | 0.003 | 0.108 | 0.979 |
| SMM | -0.040 | 0.114 | *0.725* | -0.105 | 0.138 | 0.449 |
| MM | -0.415 | 0.150 | **0.007** | -0.507 | 0.173 | **0.005** |
| Sex (Woman) | -0.078 | 0.099 | *0.433* |  |  |  |
| Age | -0.013 | 0.003 | **<0.0001** | -0.009 | 0.003 | **0.001** |
| Gammopathy subtype |  |  |  |  |  |  |
| IgG (Kappa or Lambda) | -0.104 | 0.113 | *0.362* |  |  |  |
| IgA (Kappa or Lambda) | 0.055 | 0.161 | *0.734* |  |  |  |
| IgM (Kappa or Lambda) | -0.187 | 0.198 | *0.346* |  |  |  |
| Bence Jones Kappa | -0.104 | 0.326 | *0.749* |  |  |  |
| Bence Jones Lambda | -0.331 | 0.455 | *0.453* |  |  |  |
| Immunoparesis (yes) | -0.203 | 0.097 | **0.040** | 0.043 | 0.118 | *0.721* |
| Days post-complete schedule | -0.004 | 0.001 | **<0.0001** | -0.005 | -0.005 | **<0.0001** |
| Vaccine kind (Moderna) | 0.167 | 0.102 | **0.107** | 0.258 | 0.093 | **0.007** |

Lastly, we also evaluated the impact of lymphopenia in the humoral response in patients with monoclonal gammopathies. While the total counts of lymphocytes were associated with increased levels of specific SARS-CoV-2 IgG antibodies in the univariate linear regression (estimate 0.321, *p*=0.02), the neutralization capacity was not significant (estimate 0.09, *p*=0.23).

## Discussion

In this study, we evaluated the SARS-CoV-2 vaccine-induced humoral response in different stages of the same plasma cell diseases, revealing that uninfected MM have a lower humoral response to vaccination after 3 months post-vaccine when compared with MGUS and SMM, and a group of healthy controls matched by age and sex. These results are in line with previous published data, which evaluated humoral responses at shorter time points post-SARS-CoV-2 vaccination [9–12,16,17]. Even though the MM patients showed low levels of specific SARS-CoV-2 antibodies, most of these antibodies showed to be functional demonstrated by a high ratio of neutralization/total anti-SARS-CoV-2. Despite we have included only MM patients during the first line of therapy, who may have better preserved immune function compared to more advanced stages of the disease [17], the levels of neutralizing antibodies remained very low, demonstrating that this population would benefit from a booster SARS-CoV-2 vaccine dose [18]. Importantly and similarly to other studies [12,19], we detected significantly higher plasma neutralization capacity in MM individuals who recovered from COVID-19 compared to their uninfected counterparts, highlighting that hybrid immunity elicit stronger immune responses even in this immunocompromised population [20,21]. Indeed, unvaccinated recovered COVID-19 MM patients on active treatment show already a superior antibody response compared to MM subjects after 1 month complete vaccine schedule [19].

The humoral response in MGUS and SMM patients has been less studied until now. We observed similar levels of SARS-CoV-2 antibodies and plasma neutralization capacity in MGUS and SMM patients compared to the healthy control group. Similarly, Terpos *et al*. did not identify significant differences between MGUS and controls regarding the development of neutralizing SARS-CoV-2 antibodies after 50 days from SARS-CoV-2 vaccination [10]. However, in this same study, SMM patients achieved more variable vaccine-induced responses and only 61% achieved a clinically relevant antibody response. In our study, MGUS and SMM showed comparable percentages of relevant neutralization activity compared to the control group (around 85-86% in all cases), being this proportion for SMM patients higher than previously described. The lower number of patients included in our study (N=22 *vs*. N=38) or the functional assay used may explain the differences observed between both studies.

The underlying causes for suboptimal humoral response to SARS-CoV-2 vaccine in uninfected MM patients are multifactorial, which includes disease-related immune dysregulation and the immunosuppression caused by the anti-myeloma therapies. First, myeloma cells can suppress the expansion of normal B-cells and the production of immunoglobulins after SARS-CoV-2 vaccine administration. However, only MM patients showed a suboptimal humoral immune response following vaccination, suggesting that the symptomatic disease plays a crucial role in immunosuppression while the asymptomatic disease may preserve humoral responses. Second, some anti-myeloma therapies could deplete B-cells and impair T-cell function, which may hamper also the response to vaccines with poorer neutralizing antibody and cellular immunity responses [8]. Various groups have observed an defective humoral response in patients on daratumumab-based therapies compared to other treatments [9–12], suggesting a dysfunction of the immune system and relation between plasma cells and bone marrow microenvironment. However, our study and others [17,19], with a reduced number of patients analysed in all cases, did not find this association on patients with daratumumab. Similarly to others [17], we did not find lower anti-SARS-CoV-2 specific antibodies or neutralization capacity on MM patients with previous HSCT compared to their counterparts. Additional studies with higher number of patients are required to validate these results.

Age may also contribute to a lower immune response to SARS-CoV-2 vaccines among non-cancer uninfected controls older than 65 years [22,23]. In this study, we observed that older age was negatively associated with the levels of anti-SARS-CoV-2 IgG antibodies as well as the neutralization capacity, as other groups have already highlighted [9,24]. Due to the older age of most MM patients, this parameter should be taken into account for future analysis.

Immunoparesis has been also associated with an inferior antibody response [24], and we did confirm these results in our univariate analysis, but was not confirmed in the multivariate validation. In that sense, we observed greater immunoparesis in SMM (77%) and MM (90%) patients compared to the 35% of MGUS subjects. Despite this high frequency in immunoparesis in SMM group, no significant differences in the levels of anti-SARS-CoV-2 antibodies or plasma neutralization capacity were detected between MGUS, SMM and the healthy control groups. Overall, our results suggest that other factors rather than immunoparesis may contribute to a lower response to vaccination in MM patients. In our cohort, SMM and MM presented similar defects in immune effector cells and B-cell disorders, and the only differential finding between both groups was anti-myeloma therapy and lymphopenia.

Interestingly, we observed that mRNA-1273 COVID-19 vaccine was associated to a higher levels of neutralization activity in our multivariate analysis, as previously described [9]. This result should be further investigated in larger longitudinal studies of patients with MM.

Even though an assessment of the immune response several months after vaccination has been performed, our study has some limitations. First, the relatively small number of participants, especially for the uninfected MM group, limited the multivariate analysis especially for the impact of treatment. In addition, we did not assess specific SARS-CoV-2 cellular responses after vaccination, which could also be used as a correlate of protection [25].

In conclusion, patients suffering from MGUS and SMM did not show significant differences in the plasma neutralization capacity compared to healthy controls, and booster vaccine dose should be administrated following the recommendations for the general population. Because our results indicate that MM patients in first line of therapy have a blunted antibody response after three months from complete vaccine administration, a booster vaccine dose in this vulnerable population is essential to develop an adequate and effective humoral response. In addition, larger longitudinal studies are to carefully follow up the vaccine-induced immune responses to rightly develop tailored booster campaigns in these cancer patients and to adapt their SARS-CoV-2 vaccination calendar to their immune needs.

## Data Availability

All data produced in the present study are available upon reasonable request to the authors

## Declarations

### Author’s Contributors

EA, BC, JB and MM conceived the study. MT, TP and JC determined and analysed anti-SARS-CoV-2 specific antibodies. EP, BT, SM, CR and JB determined and analysed neutralization activity. FM-l, RBABL, DP, NP, AC, RT, MF, LM and EG contribute to clinical management and sample collection. FM, VU and MM performed the statistical analysis. EA and MM analysed the data and prepared the figures. EA and MM coordinated the study, collected all clinical and laboratory data and wrote the manuscript, and FL-S corrected the final version of the manuscript. All authors discussed the results and approved the manuscript.

## Acknowledgements

We are deeply grateful to all participants who participated in this study. We also thank the technical staff of Hospital del Mar, Direcció d’Atenció Primària de la Metropolitana Nord and AIDS Figth foundation for sample collection, and staff of IrsiCaixa for sample processing (L. Ruiz, R. Ayen, L. Gomez, C. Ramirez, M. Martinez, T Puig). We thank CERCA Programme/Generalitat de Catalunya for institutional support and the Foundation Dormeur.

